# Trends and Determinants of Caesarean Section among Reproductive-age Women (2004/05-2022): A Multilevel Analysis of Demographic and Health Survey

**DOI:** 10.1101/2025.05.05.25327036

**Authors:** Elihuruma Eliufoo Stephano, Tegemea Patrick Mwalingo, Fabiola Vincent Moshi, Angelina Alphonse Joho, Shazra Kazumari, Victoria Godfrey Majengo, Mtoro J. Mtoro

## Abstract

**Background:** Caesarean section (CS) rates have risen significantly worldwide, including Tanzania, reflecting improved but uneven access to this life-saving intervention. These increasing rates in Sub-Saharan Africa call for studies to examine trends with the presence of increased maternal morbidity and mortality. We aimed to investigate the trends and determinants of CS among reproductive-age women using Tanzania Demographic and Health Survey (TDHS) data.

**Methods:** An analytical cross-sectional study using pooled data from multiple TDHS spanning from 2004/05 to 2022 was conducted. The study included 24,213 women of reproductive age, selected through a two-stage sampling method. Multilevel logistic regression, accounting for the complex survey design, was used to identify individual and community-level determinants of CS. Analyses were conducted using Stata 18.5. Adjusted odds ratio (AOR) with 95% confidence intervals (CI) were reported, and statistical significance was set at p < 0.05.

**Results:** The overall prevalence of CS was 6.8% (95% CI: 6.3-7.3), showing a significant increasing trend from 4.0% in 2004/05 to 10.7% in 2022 (p<0.001). At the individual level, older age, primary and secondary education, attending more than four antenatal care (ANC) visits, and belonging to higher wealth quintiles were associated with a higher likelihood of CS delivery. Conversely, ever being married or cohabiting was associated with lower odds of CS delivery. At the community level, women in poor communities had a lower likelihood of CS delivery, while geographical zones also showed significant variations in CS delivery likelihood.

**Conclusion:** The rising trend of CS in Tanzania, coupled with the identified individual and community-level determinants, underscores the importance of addressing disparities in access and utilization. Policies and interventions should focus on ensuring equitable access to CS based on need, considering both individual vulnerabilities and community contexts across different geographical zones.

## Background

The World Health Organization’s (WHO) recommendation that population-level cesarean section (CS) rates should ideally fall between 10-15% highlights a concerning trend of medically unnecessary procedures in some regions, while inadequate access persists in others [1]. Despite the WHO recommendations, caesarean section (CS) rates have risen dramatically worldwide, increasing from approximately 7% in 1999 to over 21% in 2023, with significant regional disparities [2]. While Latin America and the Caribbean report rates exceeding 40% in many countries, high-income regions consistently show rates between 25% and 32%. Sub-Saharan Africa (SSA) presents a complex picture of both underuse and overuse, with the rates ranging between 1.4% and 51.8% [3]. Countries like South Africa and Kenya report urban rates approaching 20%, while several West African nations maintain rates below 5% [4]. A review of literature in East Africa revealed the overall prevalence of 24% above the recommended rates with highest rates in Ethiopia (28.3%) and the lowest in Uganda, which was 11.9% [5]. CS rates have doubled over the past two decades in SSA but remain substantially lower than global averages, with considerable differences between countries and within-country variations.

This global pattern reflects not only differences in healthcare infrastructure but also changing medical practices, maternal preferences, socioeconomic factors, and shifting risk perceptions around childbirth [6]. This heterogeneity reflects profound inequities in access to emergency obstetric care, with rural-urban disparities particularly pronounced [4]. Recent studies indicate that while wealthier countries in the region are approaching or exceeding recommended levels, particularly in private facilities and urban centers, many lower-income countries continue to struggle with rates below 5%, far below what would be expected for meeting obstetric emergencies [3]. Despite the alarming increase in CS rate, a study showed that, 1 out 100 women dies after CS [7].

A recent study in Tanzania reported CS rates increased from 2% in 1996 to approximately 10.4% in 2022, with urban rates substantially higher than rural rates [8,9]. Another previous trend study reported in a single hospital study from 2000 to 2013 showed the prevalence to be 28.9% [10]. This upward trend, while representing improved access to life-saving interventions, raises questions about appropriate utilization, quality of care, and equitable distribution of services [11].

Multiple factors influence CS utilization in Tanzania and other low- and middle-income countries, operating at individual, facility, and systems levels [12]. Maternal factors include advanced maternal age, higher education, urban residence, higher socioeconomic status, and previous CS delivery [5,13]. Health system determinants encompass provider availability, facility type (public versus private), quality of antenatal care, and geographical accessibility of comprehensive emergency obstetric services [14]. Despite these influencing factors, the indications for performing a CS include a history of prior CS delivery, signs of fetal distress, extended duration of labor, unsuccessful labor induction, and abnormal fetal presentation or positioning [15]. The interplay of these factors remains inadequately explored, particularly regarding how these dynamics operate across Tanzania’s diverse geographical and socioeconomic landscape.

Most of the available research has often relied on facility-based data, which cannot capture population-level patterns as well as adequately account for women without access to facility-based care [8,13]. Despite existing research highlighting the rising prevalence of CS and its association with several factors, there is still a need to comprehensively understand the multilevel determinants influencing CS practices over time across diverse contexts. Current studies, including the recent analysis published, lack a multilevel analytical approach that integrates individual, household, and community-level factors simultaneously [8,9,13]. This research is essential to fill these gaps by employing a multilevel analysis of TDHS data spanning multiple years to establish patterns and interactions of determinants driving the CS trends. Therefore, this study aims to establish the trends and determinants of caesarean section among reproductive-age women by following a multilevel analysis of the 2005 to 2022 TDHS data. Despite the availability of prior studies [8,9,13], this research is needed to provide a more holistic, evidence-based understanding of the dynamics of CS over nearly two decades, which is vital for informed policy-making and health systems strengthening.

## Material and methods

### Study design, data source, population, and sampling procedure

This study was an analytical cross-sectional survey that utilised secondary data from the 2004/05-2022 TDHS, which conducts nationally representative population-based household surveys typically every five years. The Tanzania National Bureau of Statistics conducted the survey with the Ministries of Tanzania Mainland and Zanzibar, along with multiple stakeholders. Tanzania is one of the countries in East Africa, covering an area of 945,087 square kilometres, including 61,000 square kilometres of inland water. The 2022 census reports the country’s population at around 62 million [16].

Data for this study were obtained from the three rounds of TDHS that were conducted across all regions in Tanzania. Details in the DHS methodology have been explained elsewhere [17]. In summary, the target population for the TDHS includes women of reproductive age (15–49 years), men, children, and households across the 32 administrative regions in Tanzania. However, our analysis primarily focused on women of reproductive age. To minimize sampling errors, the country is stratified by geographic region and by urban/rural areas within each region, followed by a two-stage sampling to select a household to be surveyed. The first sampling is to select a primary sampling unit (PSU) and then select a household. PSUs are survey clusters that are usually based on census enumeration areas (EAs). A probability proportional to size is employed in each stratum to select the PSU. For each selected PSU, a complete household listing is done. This is then followed by selecting a fixed number of households to be surveyed using equal probability systematic sampling. Data were obtained from the Kids Records (KR) files of each DHS survey round, focusing on women aged 15–49 years who had a live birth within the five years preceding the survey. Records with missing information on CS delivery and duplicate entries were excluded. Each dataset was weighted appropriately, accounting for the complex survey design, and then appended to create a pooled dataset. The weighted sample sizes for each survey year were: 5,754 (2004/05), 5,509 (2010), 7,079 (2015/16), and 5,871 (2022), resulting in a total weighted sample of 24,213 women across all four survey rounds

### Study Variables

#### Dependent variable

The outcome variable was mode of delivery by CS, categorized as ‘Yes’ or ‘No’. Women who had a CS were coded as 1, while those who did not were coded as 0.

#### Independent variables

The current study examined variables at the individual and community levels based on the available data and literature [2,9,12,15]. The following variables were included under the individual level; age in years (15-24, 25-34 or 35-49), Partner’s age in years (15-24, 25-34, 35-44 or ≥45), women and partner’s education level (no formal education, primary, secondary, or higher), marital status (never married or ever married/cohabiting), literacy (illiterate or literate), wealth index (poor, middle or rich), media exposures (yes as listening to the radio, reading the newspaper or watching Television less than once a week or at least once a week or no if otherwise), household members (≤5 or ≥6), working status (working or not working), number of ANC visits (<4 or ≥4), parity (1, 2-4 or ≥5), sex of household head (male or female), visited by community healthcare worker (yes or no), visited health facility in the past 12 months (yes or no).

The following community-level variables were included: place of residence (urban or rural) and geographical zones (western, northern, central, lake, southern, eastern, or Zanzibar). Other community-level variables were derived by aggregating individual women’s characteristics at the cluster level. The level of poverty in the community was determined by the proportion of women in the poorer and poorest wealth quintiles, as indicated by the wealth index. It was classified as low (communities where <50% of women were in the poorer or poorest quintiles) or high (communities where ≥50% of women were in the poorer or poorest quintiles).

#### Data management and analysis

To address the complex survey design of the DHS, we applied individual sampling weights, primary sampling units (clusters), and strata. This adjustment accounted for the cluster sampling methodology and controlled for potential under- or over-representation of specific groups. Data cleaning, coding, and all statistical analyses were performed using STATA version 18.5 (STATA Corp, College Station, TX) [18]. Descriptive statistics, including means and standard deviations (SD) for continuous variables and frequencies and proportions for categorical variables, were used to summarize the data. Given the hierarchical nature of the DHS data, where women are nested within households, and households within clusters, a multilevel mixed-effects logistic regression model was employed. This method was essential to account for the lack of independence and equal variance typically violated in standard logistic regression. Our analysis involved four models: a null model, Model I which included individual-level factors, Model II which included community-level factors, and Model III which combined both individual and community-level factors.

#### Random effects and model fitness

To assess the variability in CS delivery across clusters, we calculated measures of variation including the Intra-class Correlation Coefficient (ICC), Median Odds Ratio (MOR), and Proportion Change in Variance (PCV), treating clusters as a random effect. The ICC, quantifying the proportion of total variance CS delivery attributable to between-cluster differences, was computed as ICC=(VC/(VC+3.29))×100. The MOR, representing the median odds ratio of CS delivery between high- and low-coverage clusters. The PCV, indicating the change in variance across models, was calculated as PCV=(Ve−Vmi)/Ve, where Ve is the variance in the null model and Vmi is the variance in subsequent models. The association between the likelihood of CS delivery and individual and community-level independent variables was estimated using fixed effects, presented as adjusted odds ratios (AORs) with 95% confidence intervals. Model comparison was performed using the deviance statistic (-2 * log-likelihood), with lower deviance indicating a better fit. Multilevel logistic regression analysis was conducted using the ‘melogit’ package in Stata. Multicollinearity among independent variables was assessed using the Variance Inflation Factor (VIF) prior to multivariable modelling. Given the DHS surveys are conducted approximately every five years in the same locations, some women might have participated in multiple rounds. To avoid potential issues with non-independent observations, our regression analysis focused solely on the most recent (2022) TDHS data. Statistical significance was set at p<0.05.

#### Ethics approval, consent to participate and publish

The study is based on the publicly available 2004/05-2022 TDHS datasets, which are accessible online and have been de-identified. The initial survey was approved by the National Institute of Medical Research Ethics Committee in Tanzania and the ICF Macro Ethics Committee in Calverton, New York. We obtained permission to use the DHS data from MEASURE Tanzania Demographic and Health Surveys after submitting a request outlining our data analysis plan. We downloaded the dataset from the DHS Program’s website upon receiving approval. Informed consent was obtained from participants before the interviews. All methods were conducted following the relevant guidelines and regulations. The consent for publication was not applicable.

## Results

### Sociodemographic characteristics

Table 1 presents the sociodemographic and community-level characteristics of women of reproductive age. Among the 24,213 women surveyed, the mean age was 29.2 years (standard deviation=7.3), and more than 40.0% were aged 25-34 years. More than half of the women had completed primary education, and a similar proportion were literate. In terms of socioeconomic status, more than one-third belonged to the poorest quintile, while nearly half were in the wealthiest quintile. Over half of the women were employed, and more than 50% had media exposure. Majority of women (>65.0%) were working and as expected, most households (>70%) were headed by men. Additionally, more than half of the women had visited a health facility in the past 12 months, and a similar proportion lived in the household with more than 6 members. On a community level, nearly more than 30.0% of women were from highly economically disadvantaged communities, and more than half lived in rural areas. (Table 1).

**Table 1:** Individual and community level characteristics among women of reproductive age in Tanzania, 2004/05-2020 (N=24,213)

| Characteristics | TDHS Year |  |  |  |
| --- | --- | --- | --- | --- |
|  | 2004/05<br>(N=5754) | 2010<br>(N=5509) | 2015/16<br>(N=7079) | 2022<br>(N=5871) |
| **Age group (years)** |  |  |  |  |
| 15-24 | 1934 (33.6) | 1719 (31.2) | 2314 (32.7) | 1985 (33.8) |
| 25-34 | 2633 (45.7) | 2389 (43.4) | 2982 (42.1) | 2592 (44.2) |
| 35-49 | 1188 (20.6) | 1402 (25.4) | 1782 (25.2) | 1294 (22) |
| Mean (±SD) | 28.9 (7.2) | 29.6 (7.4) | 29.3 (7.5) | 28.8 (7.1) |
| **Education Level** |  |  |  |  |
| No formal education | 1463 (25.4) | 1311 (23.8) | 1350 (19.1) | 1187 (20.2) |
| Primary | 3992 (69.4) | 3784 (68.7) | 4580 (64.7) | 3286 (56.0) |
| Secondary/Higher | 299 (5.2) | 414 (7.5) | 1149 (16.2) | 1397 (23.8) |
| **Marital Status** |  |  |  |  |
| Never married | 334 (5.8) | 362 (6.6) | 516 (7.3) | 484 (8.2) |
| Ever Married/Cohabiting | 5420 (94.2) | 5147 (93.4) | 6563 (92.7) | 5387 (91.8) |
| **Literacy** |  |  |  |  |
| Illiterate | 2000 (34.8) | 1906 (34.6) | 1989 (28.1) | 1510 (25.7) |
| Literate | 3754 (65.2) | 3603 (65.4) | 5089 (71.9) | 4361 (74.3) |
| **Working Status** |  |  |  |  |
| Not working | 868 (15.1) | 806 (14.6) | 1522 (21.5) | 2353 (40.1) |
| Working | 4886 (84.9) | 4703 (85.4) | 5557 (78.5) | 3518 (59.9) |
| **Media Exposure** |  |  |  |  |
| No | 1305 (22.7) | 1429 (25.9) | 1253 (17.7) | 2094 (35.7) |
| Yes | 4449 (77.3) | 4080 (74.1) | 5826 (82.3) | 3777 (64.3) |
| **ANC Visits** |  |  |  |  |
| <4 | 2217 (38.6) | 3134 (57.0) | 3490 (49.3) | 2049 (34.9) |
| ≥4 | 3532 (61.4) | 2361 (43.0) | 3589 (50.7) | 3822 (65.1) |
| **Parity** |  |  |  |  |
| 1 | 1170 (20.3) | 1057 (19.1) | 1730 (24.4) | 1310 (22.3) |
| 2-4 | 2798 (48.6) | 2694 (48.9) | 3285 (46.4) | 3065 (52.2) |
| ≥5 | 1286 (31.1) | 1764 (32.0) | 2063 (29.2) | 1496 (25.5) |
| **Wealth Index** |  |  |  |  |
| Poorest | 2409 (41.9) | 2305 (41.8) | 2947 (41.6) | 2437 (41.5) |
| Middle | 1166 (20.3) | 1171 (21.3) | 1349 (19.1) | 1140 (19.4) |
| Rich | 2179 (37.8) | 2034 (36.9) | 2783 (39.3) | 2294 (39.1) |
| **Sex of Household Head** |  |  |  |  |
| Male | 4676 (81.3) | 4481 (81.3) | 5751 (81.2) | 4586 (78.1) |
| Female | 1078 (18.7) | 1028 (18.7) | 1327 (18.8) | 1285 (21.9) |
| **Visited Health facility in the past 12 months** |  |  |  |  |
| No | 1810 (31.5) | 1277 (23.2) | 1689 (23.9) | 1842 (31.4) |
| Yes | 3944 (68.5) | 4232 (76.8) | 5389 (76.1) | 4029 (68.6) |
| **Household members** |  |  |  |  |
| ≤5 | 2581 (44.9) | 2361 (42.9) | 3053 (43.1) | 2837 (48.3) |
| ≥6 | 3173 (55.1) | 3148 (57.1) | 4025 (56.9) | 3034 (51.7) |
| **Partner age group (years)** |  |  |  |  |
| 15-24 | 357 (7.4) | 300 (6.7) | 436 (7.7) | 380 (7.9) |
| 25-34 | 2048 (42.2) | 1774 (39.4) | 2167 (38.1) | 2048 (42.4) |
| 35-44 | 1524 (31.4) | 1545 (34.3) | 2022 (35.6) | 1580 (32.7) |
| 45+ | 923 (19) | 882 (19.6) | 1059 (18.6) | 821 (17) |
| Mean (±SD) | 36.8 (10.2) | 37.4 (9.9) | 36.7 (9.6) | 36.1 (9.5) |
| **Partner education** |  |  |  |  |
| No formal education | 965 (17.9) | 870 (17.0) | 752 (13.2) | 676 (14.0) |
| Primary | 3963 (73.3) | 3741 (72.9) | 3906 (68.7) | 2921 (60.7) |
| Secondary/Higher | 477 (8.8) | 519 (10.1) | 1025 (18.1) | 1217 (25.3) |
| **Community poverty** |  |  |  |  |
| Low | 3569 (62.0) | 3377 (61.3) | 4396 (62.1) | 3668 (62.5) |
| High | 2186 (38.0) | 2133 (38.7) | 2683 (37.9) | 2204 (37.5) |
| **Residence** |  |  |  |  |
| Urban | 1267 (22.0) | 1270 (23.1) | 2123 (30.0) | 1648 (28.1) |
| Rural | 4487 (78.0) | 4239 (76.9) | 4955 (70.0) | 4223 (71.9) |
| **Geographical Zones** |  |  |  |  |
| Western | 592 (10.3) | 549 (10) | 779 (11) | 597 (10.2) |
| Northern | 600 (10.4) | 648 (11.8) | 699 (9.9) | 646 (11.0) |
| Central | 646 (11.2) | 634 (11.5) | 795 (11.2) | 586 (10.0) |
| Southern | 1347 (23.4) | 1272 (23.1) | 1482 (20.9) | 1130 (19.3) |
| Lake | 1669 (29) | 1522 (27.6) | 2015 (28.5) | 1936 (33) |
| Eastern | 757 (13.1) | 743 (13.5) | 1137 (16.1) | 807 (13.7) |
| Zanzibar | 144 (2.5) | 141 (2.6) | 171 (2.4) | 170 (2.9) |

ANC; Antenatal care, SD; Standard deviation

### Prevalence and trend in cesarean section delivery

The overall prevalence of CS delivery was 6.8% (95% CI: 6.3-7.3). Examining the trend over time, the prevalence significantly increased linearly (p<0.001) from 4.0% (95% CI: 3.4-4.9) in 2004/05 to 5.4% (95% CI: 4.6-6.3) in 2010, 6.9% (95% CI: 6.1-7.9) in 2015/16, and 10.7% (95% CI: 9.5-11.9) in 2022. (Figure 1).

**Figure 1.**
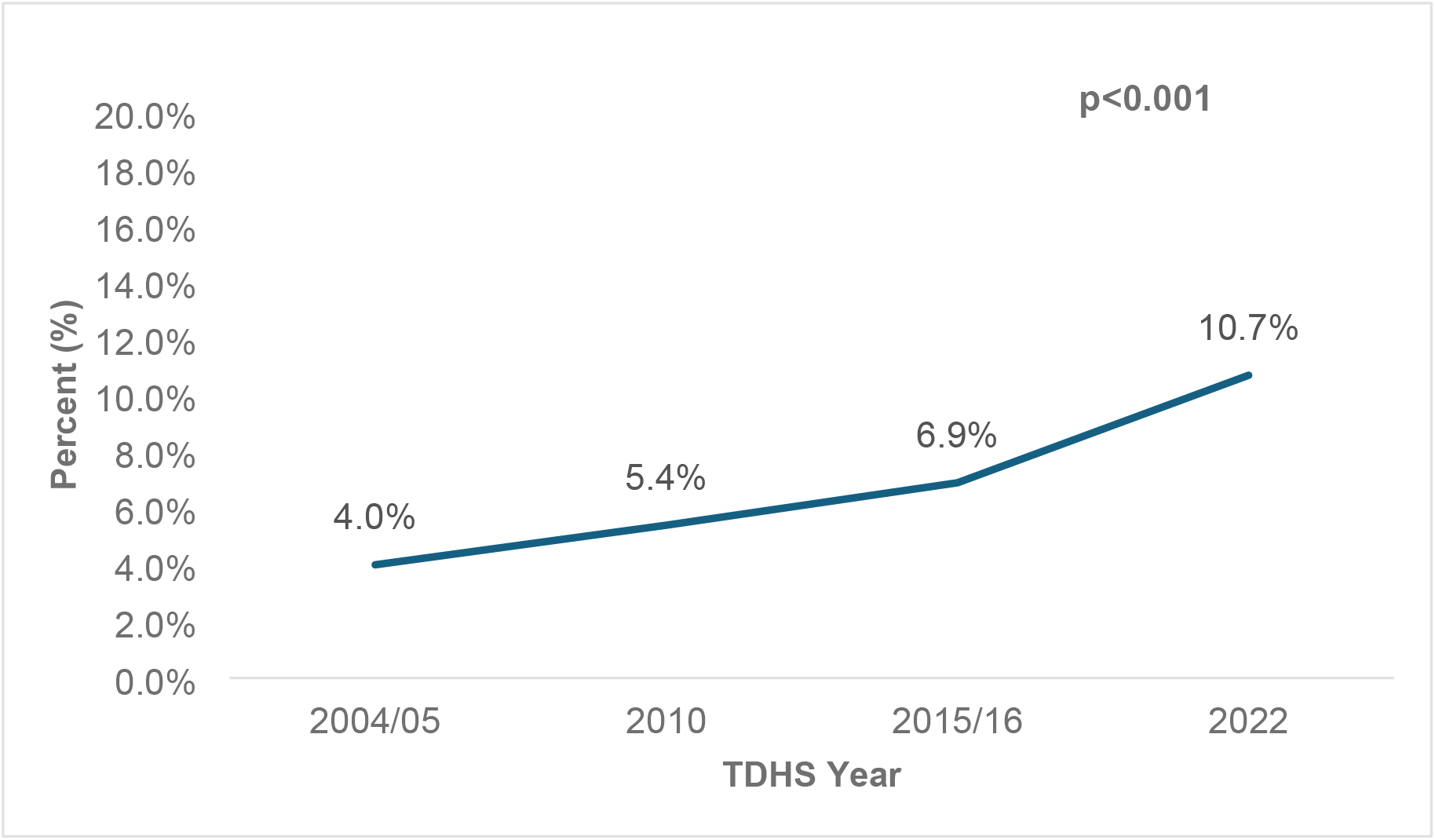
Trend in caesarian section delivery among women of reproductive age in Tanzania (2004/05-2022)

### Distribution of caesarian section delivery by sociodemographic characteristics

The bivariate analysis consistently demonstrates significant associations (p<0.05) between several socio-economic and demographic factors and CS delivery across the four survey years (2004/05, 2010, 2015/16, and 2022). Specifically, literacy, education level, literacy, media exposure, wealth index, parity, partner education, residence, and geographical zone appeared to be important correlates of CS delivery among women.

Notably, CS delivery remained persistently high among women with secondary/higher education rising from 11.9% in 2004/05 to 19.6% in 2022. Conversely, the percentage of CS delivery was notably low among women with no formal education with the highest (4.3%) being reported in 2022. Additionally, literate women had higher proportion of CS delivery raised form 4.9% in 2004/05 to 12.8% in 2022.

Media exposure demonstrated a significant association (p<0.001) across the four survey years. The proportion of CS delivery increased from 4.6% in 2004/05 to 12.9% in 2022 among women with media exposure. Women in the wealthiest households demonstrated a higher proportion of CS delivery over time, from 6.0% in 2004/05 to 16.4% in 2020. On the other hand, women with one child demonstrated a higher proportion of CS delivery compared to their counterparts. In terms of geographical location, urban settings consistently displayed a higher proportion of CS delivery compared to rural areas, with the proportion increasing linearly over time from 8.1% in 2004/05 to 17.4% in 2022. The Northern, Central, Southern, and Eastern zones of mainland Tanzania consistently showed higher prevalence rates of CS delivery in the survey years, while the Lake zone consistently exhibited a notably lower proportion of CS delivery (Table 2).

**Table 2:** Distribution of caesarian section delivery by sociodemographic characteristics among women of reproductive age in Tanzania, 2004/05-2020 (N=24,213)

| Characteristics | TDHS Year |  |  |  |
| --- | --- | --- | --- | --- |
|  | 2004/05<br>% [95%CI] | 2010<br>% [95%CI] | 2015/16<br>% [95%CI] | 2022<br>% [95%CI] |
| **Age group (years)** | ns | ns | * | ns |
| 15-24 | 4.4 [3.4-5.7] | 5.2 [4.0-6.7] | 5.0 [4.0-6.1] | 10.0 [6.2-12.2] |
| 25-34 | 4.2 [3.3-5.5] | 5.8 [4.7-7.2] | 6.3 [6.9-10.0] | 10.7 [9.2-12.4] |
| 35-49 | 3.0 [1.9-4.6] | 5.1 [3.9-6.6] | 7.3 [5.8-9.1] | 11.6 [9.5-13.9] |
| **Education Level** | *** | *** | *** | *** |
| No formal education | 1.6 [1.0-2.6] | 2.4 [1.6-3.8] | 2.9 [2.0-4.1] | 4.3 [2.6-6.9] |
| Primary | 4.3 [3.6-5.2] | 5.4 [3.6-6.3] | 5.9 [5.3-6.9] | 9.2 [8.0-10.6] |
| Secondary/Higher | 11.9 [6.8-19.8] | 15.1 [11.1-20.0] | 15.9 [13.0-19.4] | 19.6 [16.9-22.5] |
| **Marital Status** | *** | * | ns | * |
| Never married | 8.0 [5.2-12.1] | 8.9 [5.9-12.9] | 8.5 [6.1-11.7] | 17.2 [13.5-21.6] |
| Ever Married/Cohabiting | 3.8 [3.1-4.6] | 5.2 [4.4-6.0] | 6.8 [6.0-7.8] | 10.1 [8.8-11.4] |
| **Literacy** | *** | * | *** | *** |
| Illiterate | 2.3 [1.7-3.3] | 3.9 [2.8-5.3] | 3.2 [2.4-4.2] | 4.4 [3.1-6.1] |
| Literate | 4.9 [4.0-6.0] | 6.2 [5.3-7.3] | 8.4 [7.4-9.7] | 12.8 [11.4-14.3] |
| **Working Status** | ns | ns | ns | *** |
| Not working | 5.5 [3.6-8.3] | 5.6 [3.9-7.9] | 6.4 [5.1-8.0] | 8.9 [7.5-10.6] |
| Working | 3.8 [3.1-4.6] | 5.4 [4.5-6.3] | 7.1 [6.2-8.2] | 11.8 [10.3-13.6] |
| **Media Exposure** | *** | *** | *** | *** |
| No | 2.1 [1.4-3.2] | 2.9 [2.0-4.1] | 3.5 [2.4-5.0] | 6.6 [5.4-8.0] |
| Yes | 4.6 [3.8-5.6] | 6.3 [5.3-7.4] | 7.7 [6.8-8.8] | 12.9 [11.3-14.6] |
| **ANC Visits** | *** | *** | *** | *** |
| <4 | 2.6 [1.9-3.6] | 4.3 [3.4-5.3] | 5.0 [4.1-6.0] | 6.8 [5.5-8.3] |
| ≥4 | 5.0 [4.1-6.0] | 7.0 [5.7-8.6] | 8.9 [7.7-10.3] | 12.7 [11.2-14.4] |
| **Parity** | *** | *** | *** | *** |
| One | 6.2 [4.6-8.3] | 9.1 [7.1-11.5] | 10.1 [8.5-12.1] | 16.4 [13.5-19.5] |
| 2-4 | 4.5 [3.5-5.6] | 5.4 [4.4-6.6] | 7.8 [6.5-9.3] | 10.7 [9.4-12.3] |
| ≥5 | 2.0 [1.3-3.0] | 3.3 [2.5-4.5] | 3.0 [2.3-4.0] | 5.5 [4.1-7.1] |
| **Wealth Index** | *** | *** | *** | *** |
| Poor | 2.3 [1.7-3.1] | 2.7 [2.0-3.6] | 3.1 [2.4-4.0] | 5.4 [4.4-6.5] |
| Middle | 3.9 [2.8-5.6] | 4.2 [2.9-5.8] | 4.7 [3.5-6.4] | 10.4 [8.3-13.0] |
| Rich | 6.0 [4.8-7.6] | 9.1 [7.6-10.9] | 12.2 [10.5-14.0] | 16.4 [13.9-19.2] |
| **Sex of Household Head** | ns | ns | * | ns |
| Male | 3.9 [3.2-4.8] | 5.5 [4.6-6.4] | 6.4 [5.8-7.4] | 10.6 [9.2-12.0] |
| Female | 5.7 [3.3-6.5] | 5.1 [3.7-7.1] | 8.9 [6.8-11.7] | 10.9 [9.1-13.2] |
| **Visited Health facility in the past 12 months** | ns | ns | ns | *** |
| No | 3.3 [2.4-4.5] | 5.7 [4.4-7.5] | 5.9 [4.5-7.8] | 7.2 [6.0-8.8] |
| Yes | 4.4 [3.6-5.4] | 5.3 [4.5-6.3] | 7.3 [6.4-8.3] | 12.2 [10.7-13.9] |
| **Household members** | ns | ns | * | *** |
| ≤5 | 4.4 [3.5-5.7] | 6.2 [5.0-7.5] | 7.9 [6.7-9.2] | 12.8 [11.2-14.5] |
| ≥6 | 3.7 [2.9-4.7] | 4.9 [3.9-5.9] | 6.3 [5.3-7.4] | 8.6 [7.3-10.2] |
| **Partner age group (years)** | ns | ns | ns | ns |
| 15-24 | 3.5 [1.9-6.4] | 6.1 [3.4-10.7] | 5.9 [3.6-9.6] | 11.4 [6.9-18.2] |
| 25-34 | 4.6 [3.5-5.9] | 5.9 [4.7-7.3] | 6.6 [5.4-8.0] | 10.7 [9.1-12.5] |
| 35-44 | 3.2 [2.0-4.9] | 5.0 [3.9-6.6] | 7.6 [6.3-9.3] | 10.1 [8.2-12.4] |
| 45+ | 2.9 [1.8-4.7] | 4.4 [3.0-6.3] | 6.9 [5.3-9.1] | 9.1 [6.9-11.9] |
| **Partner education** | *** | *** | *** | *** |
| No formal education | 1.1 [0.6-2.0] | 1.9 [1.1-3.3] | 2.8 [1.6-4.8] | 5.1 [3.4-7.5] |
| Primary | 3.8 [3.1-4.7] | 4.9 [4.1-5.9] | 5.8 [4.9-6.8] | 8.5 [7.3-10.1] |
| Secondary/Higher | 9.0 [5.6-14.3] | 12.6 [9.6-16.4] | 14.7 [11.9-18.0] | 17.3 [14.6-20.3] |
| **Residence** | *** | *** | *** | *** |
| Urban | 8.1 [6.1-10.6] | 10.1 [8.0-12.7] | 12.6 [10.5-15.1] | 17.4 [14.2-21.1] |
| Rural | 2.9 [2.4-3.6] | 4.0 [3.3-4.8] | 4.5 [3.8-5.4] | 8.0 [6.9-9.2] |
| **Geographical Zones** | * | *** | *** | *** |
| Western | 3.0 [1.6-5.7] | 4.5 [2.6-7.5] | 4.3 [2.9-6.3] | 6.5 [4.4-9.6] |
| Northern | 4.4 [2.9-6.6] | 7.3 [5.1-10.2] | 9.2 [6.7-12.6] | 14.8 [10.7-20.0] |
| Central | 2.4 [1.4-3.9] | 3.4 [2.3-5.0] | 5.6 [4.1-7.7] | 12.2 [8.5-17.2] |
| Southern | 3.8 [2.7-5.2] | 6.6 [4.9-8.7] | 8.5 [7.1-10.3] | 14.8 [12.7-17.2] |
| Lake | 3.7 [2.6-5.3] | 3.2 [2.1-5.0] | 3.6 [2.6-4.9] | 4.4 [3.4-5.9] |
| Eastern | 7.4 [4.6-11.7] | 8.6 [6.0-12.2] | 12.2 [9.1-16.4] | 17.5 [12.7-23.7] |
| Zanzibar | 2.5 [1.6-3.9] | 5.9 [4.3-7.9] | 7.0 [5.8-8.4] | 14.2 [10.4-19.3] |

\*\*\*p<0.001; \*p<0.05; ns; not significant; CI: Confidence Interval

### Multilevel analysis of individual and community determinants of caesarian section delivery

Table 3 presents multilevel analysis findings, Women aged 35-49 (AOR=1.38, 95%CI: 1.07-1.77) had higher likelihood of CS delivery compared to women aged 15-24. In terms of education, women with primary (AOR=2.01, 95%CI:1.42-2.84) and secondary/higher (AOR=3.40, 95%CI:2.32-4.96) had increased odds of CS delivery compared to women with no formal education. Women with more than four ANC visited had higher likelihood of CS delivery compared to their counterparts (AOR=1.33, 95%CI:1.08-1.63). At the community level, women from poorer community had significantly lower likelihood of CS delivery compared to their counterparts (AOR=0.66, 95%CI:0.49-0.90). Geographical zones in the mainland, Northern (AOR=2.59, 95%CI:1.76-3.81), Central (AOR=2.03, 95%CI:1.32-3.14), Southern (AOR=2.34, 95%CI:1.69-3.25) and Eastern (AOR=2.06, 95%CI:1.40-3.05) had had increased odds of CS delivery compared to Zanzibar (Table 3).

**Table 3:**
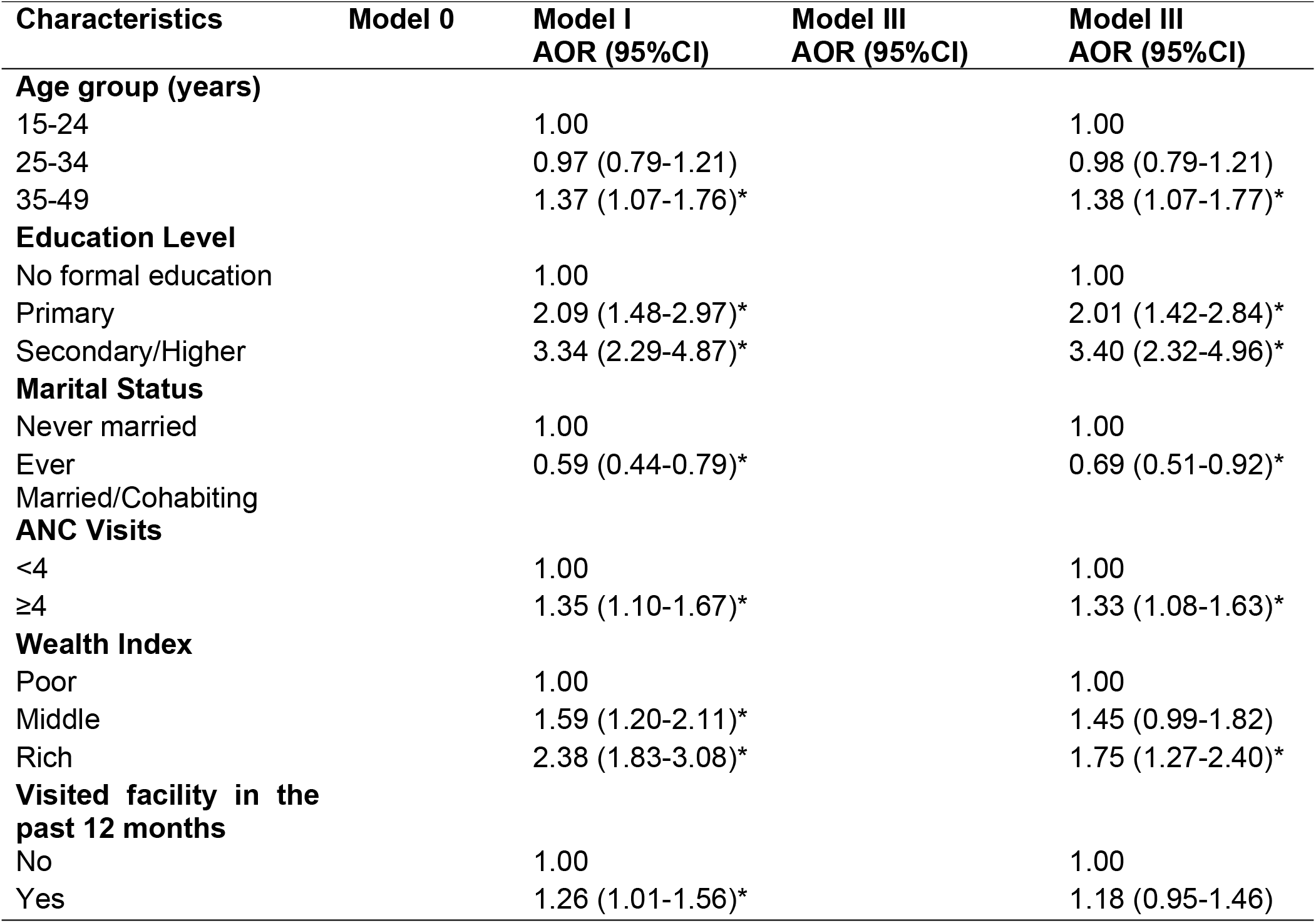
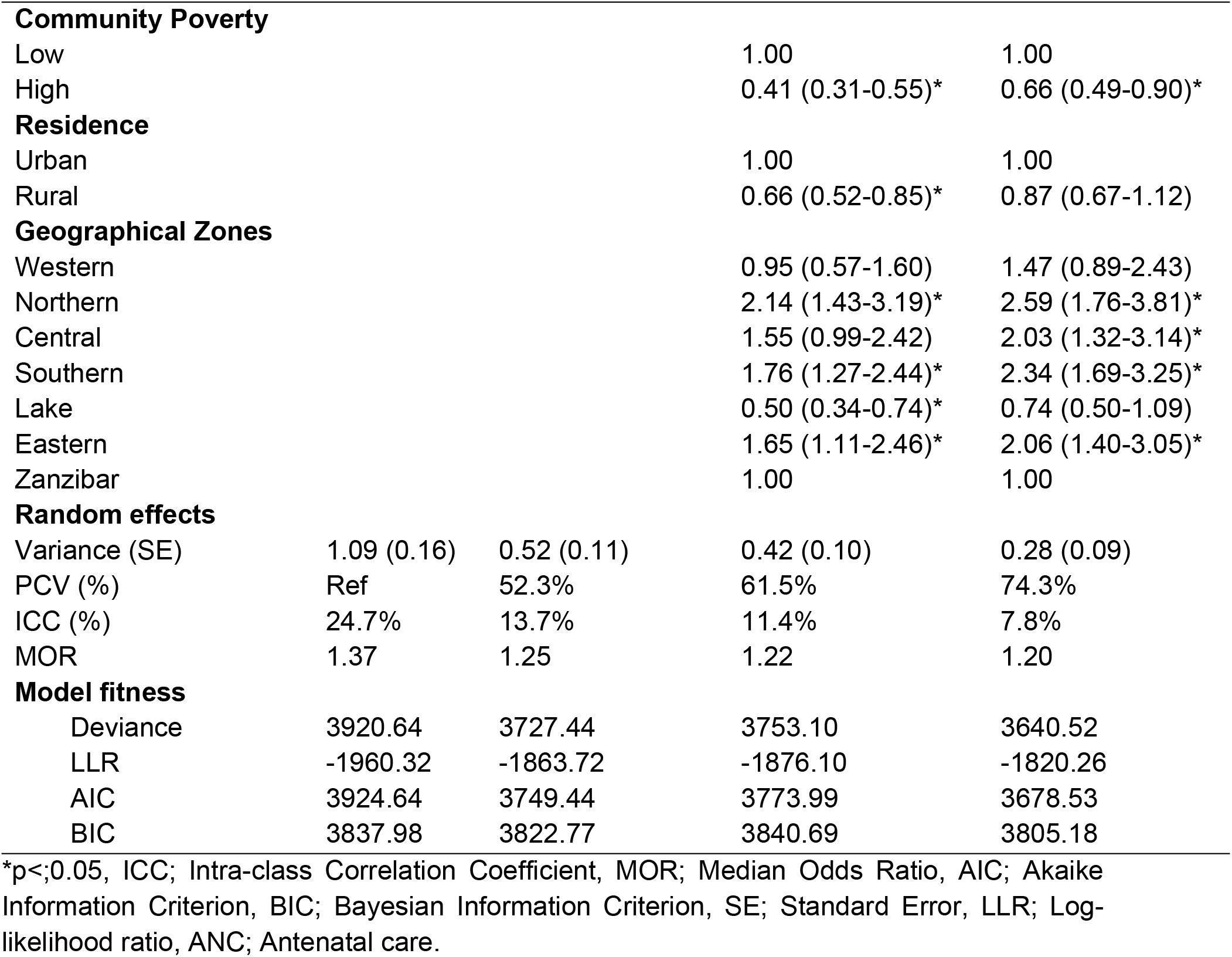
Multilevel Mixed-effects Logistic regression model for determinants of caesarian section delivery among women of reproductive age in Tanzania, DHS 2022 (N=5,871)

| Characteristics | Model 0 | Model I<br>AOR (95%CI) | Model III<br>AOR (95%CI) | Model III<br>AOR (95%CI) |
| --- | --- | --- | --- | --- |
| **Age group (years)** |  |  |  |  |
| 15-24 |  | 1.00 |  | 1.00 |
| 25-34 |  | 0.97 (0.79-1.21) |  | 0.98 (0.79-1.21) |
| 35-49 |  | 1.37 (1.07-1.76)* |  | 1.38 (1.07-1.77)* |
| **Education Level** |  |  |  |  |
| No formal education |  | 1.00 |  | 1.00 |
| Primary |  | 2.09 (1.48-2.97)* |  | 2.01 (1.42-2.84)* |
| Secondary/Higher |  | 3.34 (2.29-4.87)* |  | 3.40 (2.32-4.96)* |
| **Marital Status** |  |  |  |  |
| Never married |  | 1.00 |  | 1.00 |
| Ever<br>Married/Cohabiting |  | 0.59 (0.44-0.79)* |  | 0.69 (0.51-0.92)* |
| **ANC Visits** |  |  |  |  |
| <4 |  | 1.00 |  | 1.00 |
| ≥4 |  | 1.35 (1.10-1.67)* |  | 1.33 (1.08-1.63)* |
| **Wealth Index** |  |  |  |  |
| Poor |  | 1.00 |  | 1.00 |
| Middle |  | 1.59 (1.20-2.11)* |  | 1.45 (0.99-1.82) |
| Rich |  | 2.38 (1.83-3.08)* |  | 1.75 (1.27-2.40)* |
| **Visited facility in the**<br>**past 12 months** |  |  |  |  |
| No |  | 1.00 |  | 1.00 |
| Yes |  | 1.26 (1.01-1.56)* |  | 1.18 (0.95-1.46) |
| **Community Poverty** |  |  |  |  |
| Low |  |  | 1.00 | 1.00 |
| High |  |  | 0.41 (0.31-0.55)* | 0.66 (0.49-0.90)* |
| **Residence** |  |  |  |  |
| Urban |  |  | 1.00 | 1.00 |
| Rural |  |  | 0.66 (0.52-0.85)* | 0.87 (0.67-1.12) |
| **Geographical Zones** |  |  |  |  |
| Western |  |  | 0.95 (0.57-1.60) | 1.47 (0.89-2.43) |
| Northern |  |  | 2.14 (1.43-3.19)* | 2.59 (1.76-3.81)* |
| Central |  |  | 1.55 (0.99-2.42) | 2.03 (1.32-3.14)* |
| Southern |  |  | 1.76 (1.27-2.44)* | 2.34 (1.69-3.25)* |
| Lake |  |  | 0.50 (0.34-0.74)* | 0.74 (0.50-1.09) |
| Eastern |  |  | 1.65 (1.11-2.46)* | 2.06 (1.40-3.05)* |
| Zanzibar |  |  | 1.00 | 1.00 |
| **Random effects** |  |  |  |  |
| Variance (SE) | 1.09 (0.16) | 0.52 (0.11) | 0.42 (0.10) | 0.28 (0.09) |
| PCV (%) | Ref | 52.3% | 61.5% | 74.3% |
| ICC (%) | 24.7% | 13.7% | 11.4% | 7.8% |
| MOR | 1.37 | 1.25 | 1.22 | 1.20 |
| **Model fitness** |  |  |  |  |
| Deviance | 3920.64 | 3727.44 | 3753.10 | 3640.52 |
| LLR | -1960.32 | -1863.72 | -1876.10 | -1820.26 |
| AIC | 3924.64 | 3749.44 | 3773.99 | 3678.53 |
| BIC | 3837.98 | 3822.77 | 3840.69 | 3805.18 |

\*p<;0.05, ICC; Intra-class Correlation Coefficient, MOR; Median Odds Ratio, AIC; Akaike Information Criterion, BIC; Bayesian Information Criterion, SE; Standard Error, LLR; Log-likelihood ratio, ANC; Antenatal care.

### Measures of variation and model fitness

A null model was used to assess community-level variation in CS delivery and reveal significant differences across localities (variance = 1.09, p < 0.001), with 24.7% of the total variation between clusters and 75.3% within clusters. The null model’s Median Odds Ratio (MOR) of 1.37 indicated substantial community-level effects. Model I (individual factors) showed 13.7% individual-level variation (MOR = 1.25), while Model II (community factors) had an MOR of 1.25 and an Intraclass Correlation Coefficient (ICC) of 11.4%, indicating cluster-level variation. The final model (Model III), incorporating both levels, demonstrated the best fit (lowest deviance, highest LLR), with 7.8% of the variation attributed to both individual and community factors and an MOR of 1.20. As detailed in Table 3, these findings highlight the significant influence of both individual and community-level factors on CS delivery among Tanzanian women of reproductive age.

## Discussion

This study assessed the trends and determinants of CS delivery among reproductive-age women in Tanzania, utilizing nationally representative data from four survey waves (2004/05-2022). The study findings reveal a clear upward trend in CS rates, rising from 4% in 2004/05 to 10.7% in 2022, with significant variations across individual demographics, socioeconomic factors, and community-level factors.

The steady rise in Tanzania’s CS rate reflects important progress in expanding access to emergency obstetric care while remaining within the WHO-recommended optimal CS population rate [19]. This trajectory demonstrates Tanzania’s success in gradually improving the availability of life-saving surgical interventions without exceeding evidence-based thresholds, which is a notable achievement compared to many countries experiencing rapid overmedicalization of CS birth [3]. The most significant acceleration occurred between 2015 and 2022 (6.9% to 10.7%), likely reflecting targeted health system investments during Tanzania’s One Plan II implementation period [20]. Tanzania’s increasing trend in CS use mirrors Rwanda’s, though at a slower pace. In contrast to Tanzania’s gradual rise, Rwanda experienced a much sharper increase, a more than fivefold surge [21]. While both Tanzania and Rwanda have seen significant rises in CS rates, Rwanda’s rapid increase-reaching the upper threshold of WHO’s recommended range, calls for careful monitoring to prevent overuse and ensure procedures remain medically justified [21]. Tanzania’s slower growth suggests room for targeted improvements in access, particularly in rural areas, while guarding against unnecessary interventions. Both countries must balance equitable availability of life-saving CS with evidence-based practices to optimize maternal and neonatal health outcomes.

Additionally, Tanzania’s rise in cesarean deliveries reflects broader global and regional patterns, yet with critical distinctions. Globally, CS rates averaged 21.1% in 2018 [3], far exceeding Tanzania’s current rate, with high-income countries driving the overuse of C-sections [2]. Tanzania’s national CS rate (10.7%) has surpassed the SSA regional average of 5% [3], reflecting progress in obstetric care availability. However, this aggregate figure obscures a critical double burden of inequalities (dangerous underuse among marginalized groups and potential overuse among the advantaged population) rooted in socioeconomic and geographic barriers. Striking disparities observed in our analysis reveal this dichotomy.

For instance, women who have secondary or higher education are accessing C-section at 19.6% compared to just 4.3% for those with no formal education, which is a more than fourfold difference, suggesting potential overuse of CS amongst the educated group and access below the life-saving threshold in the uneducated group. This disparity mirrors patterns observed in other low-income countries [22,23]. This pattern repeats across several socioeconomic demographics. Such extremes suggest that Tanzania faces simultaneous crises of inequitable access and questionable overmedicalization of births.

The overuse among privileged groups, evidenced by rates exceeding 15% among educated, richest women, urban residents, and wealthy women, may reflect non-medical factors such as provider preferences and convenience, financial incentives, maternal requests, or too much too soon interventions [24]. Conversely, the underuse among disadvantaged women, particularly those with no education, rural residents, and in the Western zone Tanzania signals systemic failures in equitable service delivery [25]. These disparities violate the WHO’s principle of appropriate use [19]. These observations underscore the need for policy actions to address both sides of the disparities. Without such dual-focused interventions, Tanzania risks perpetuating a cycle where overuse for some coexists with deadly shortage for others, a paradox undermining progress toward maternal health equity [25].

### Determinants of CS in Tanzania

Multivariable analysis identified several significant individual and community-level determinants of cesarean delivery in Tanzania. Older age (35-49 years) was associated with increasing cesarean delivery compared to younger women (15-24 years). This finding is consistent with the pattern of increased CS rate with advanced maternal age observed amongst Danish births [26] and the global pattern of CS and maternal age in the first childbearing [27]. Educational attainment showed a particularly strong gradient, with women having primary education and secondary/higher education demonstrating substantially higher odds compared to those with no formal education. These findings suggest their independent role in shaping access to surgical obstetric care. The mechanisms underlying this relationship are multifactorial whereby some researchers indicated that higher education levels are associated with greater literacy, enabling more informed demand for services [25], better ability to navigate complex health systems [28], and increased financial resources to access private facilities where cesarean section rates are typically higher [29]. However, the magnitude of these disparities in Tanzania exceeds those reported in comparable low-income settings [24], indicating particularly entrenched inequalities in service provision. These findings underscore the need for interventions that address both supply-side barriers, improving equitable access to emergency obstetric care, and demand-side factors to mitigate education-based disparities in maternal healthcare utilization.

Antenatal care (ANC) visits emerged as another significant predictor. The association between higher ANC attendance and increased cesarean rates warrants careful consideration, provided Tanzania’s ANC coverage patterns. While 65% of pregnant women complete the recommended four or more ANC visits, this still leaves 35% without adequate antenatal care [17]. The positive relationship between ANC attendance and CS delivery may suggest two potential scenarios including appropriate identification of high-risk pregnancies requiring intervention among women receiving comprehensive ANC attendance, and potential overmedicalization of care for this group, particularly given the strong correlation between ANC completion and higher education levels of cesarean delivery. Meanwhile, the remaining women with fewer ANC visits may face both under-detection of true obstetric risks and systemic barriers to accessing timely cesarean delivery when medically needed [24].

Geographic disparities were particularly striking, with all mainland zones showing significantly higher odds of CS compared to Zanzibar. The Northern zone had the highest odds, followed by the Southern, the Eastern, and the Central zones. These findings align consistently with previous studies documenting Zanzibar’s unusually low cesarean rates [30]. While there has been a consistent documentation of CS disparity between Zanzibar and the Tanzania mainland, peer-reviewed studies specifically investigating Zanzibar’s lower CS rates and geographical zone CS distributions are notably absent. This evidence gap warrants urgent research attention that will guide the design of the contextualized and tailored interventions for addressing this disparity.

### Strengths and limitations

This study possesses several notable strengths. Firstly, the use of nationally representative data spanning from the 2004/05-2022 TDHS, combined with a large sample size, enhances the generalizability of our findings. Secondly, the rigorous methodological procedures employed during data collection, including the involvement of highly experienced field assistants, contribute to the high quality of the data. Finally, the use of multilevel binary logistic regression enabled a robust analysis and addressed potential validity concerns. However, the study’s findings should be interpreted with consideration of its limitations. The cross-sectional design of the TDHS data precludes the establishment of causal relationships. Due to the self-reported nature of many variables, recall and social desirability biases may be present.

### Implications for Practice and Policy Recommendations

Healthcare practices should address the identified disparities in CS delivery linked to individual characteristics (age, education, wealth, marital status, ANC attendance) and community factors (community wealth, geographical zone). Health providers need to be vigilant in identifying women at risk of poor maternal outcomes. A key focus should be improving access to comprehensive emergency obstetric care, including CS, in underserved, poorer communities through stronger referral systems, reduced financial obstacles, and ensuring adequate resources and skilled personnel. Conversely, practices serving higher socioeconomic groups should be reviewed to guarantee CS is performed based on medical need, guided by evidence-based guidelines, and shared decision-making with patients. ANC offers a vital platform for risk assessment, counselling on delivery options, and timely referrals. Therefore, pregnant women should be emphasized on the significance of ANC visits.

Policy interventions are crucial to ensure equitable access to CS. Financial barriers, particularly for low-income women, should be reduced through mechanisms like subsidies or insurance. National, evidence-based guidelines for appropriate CS indications are necessary to promote consistent and rational utilization across all healthcare facilities, minimizing unwarranted variations.

Furthermore, public health education campaigns are needed to enhance awareness regarding the importance of timely access to appropriate obstetric care, including when a CS is medically indicated, while also addressing potential misconceptions or preferences for CS without medical necessity. A comprehensive approach integrating these practices and policy changes is vital for optimizing CS utilization in Tanzania.

## Conclusion

Tanzania’s rising CS rates, while signifying advances in emergency obstetric care access, simultaneously illuminate profound and persistent inequities entrenched within the healthcare system that threaten maternal health equity. The coexistence of underuse among disadvantaged populations and potential overuse within the privileged underscores a complex paradox requiring nuanced health policy responses that transcend simple supply expansion. Equitable improvement demands targeted strategies that dismantle geographic and socioeconomic barriers, particularly in rural and Zanzibar regions, while instituting the best stewardship to prevent unnecessary interventions driven by non-medical factors. Enhancing antenatal care quality and ensuring its role as a precise risk-assessment tool, rather than a pathway to overmedicalization, will be critical, alongside community-tailored research to understand and address Zanzibar’s distinct context. Ultimately, fostering a health system that balances access with appropriate, evidence-based caesarean utilization is essential for safeguarding maternal and neonatal outcomes, demanding integrated policy interventions that address both demand- and supply-side challenges within Tanzania’s evolving obstetric landscape.

## Data Availability

The raw data supporting the conclusions of this article will be made available by the authors without undue reservation. The complete dataset is available at https://dhsprogram.com.

## Acknowledgements

We thank the DHS program for making the data available for this study, and TILAM International for statistical consultation.

## Authors’ Contribution

EES and MJM conceptualized the idea and conducted the formal analysis. EES, TPM, FVM, AAJ, SK, VGM and MJM, supported interpretation, manuscript preparation and reviewed subsequent versions of the manuscript. All authors read and approved the final manuscript.

## Funding

Not Applicable.

## Competing interests

The authors declare that the research was conducted without any commercial or financial relationships that could be construed as a potential conflict of interest.

## Notes

### Competing Interest Statement

The authors have declared no competing interest.

### Funding Statement

No funding

## Reference

1. Betran A, Torloni M, Zhang J, Gülmezoglu A, Aleem H, Althabe F, et al. WHO Statement on Caesarean Section Rates. BJOG. 2016;123:667–70.

2. Angolile CM, Max BL, Mushemba J, Mashauri HL. Global increased cesarean section rates and public health implications: A call to action. Health Sci Rep. 2023;6:e1274.

3. Betran AP, Ye J, Moller A-B, Souza JP, Zhang J. Trends and projections of caesarean section rates: global and regional estimates. BMJ Glob Health. 2021;6:e005671.

4. Ahinkorah BO, Aboagye RG, Seidu A-A, Okyere J, Mohammed A, Chattu VK, et al. Rural–urban disparities in caesarean deliveries in sub-Saharan Africa: a multivariate non-linear decomposition modelling of Demographic and Health Survey data. BMC Pregnancy and Childbirth. 2022;22:709.

5. Habteyes AT, Mekuria MD, Negeri HA, Kassa RT, Deribe LK, Sendo EG. Prevalence and associated factors of caesarean section among mothers who gave birth across Eastern Africa countries: Systematic review and meta-analysis study. Heliyon [Internet]. 2024 [cited 2025 Apr 18];10. Available from: https://www.cell.com/heliyon/abstract/S2405-8440(24)08542-6

6. Yihune Teshale M, Bante A, Gedefaw Belete A, Crutzen R, Spigt M, Stutterheim SE. Barriers and facilitators to maternal healthcare in East Africa: a systematic review and qualitative synthesis of perspectives from women, their families, healthcare providers, and key stakeholders. BMC Pregnancy and Childbirth. 2025;25:111.

7. Mose A, Abebe H. Magnitude and associated factors of caesarean section deliveries among women who gave birth in Southwest Ethiopia: institutional-based cross-sectional study. Archives of Public Health. 2021;79:1–9.

8. Cavallaro FL, Pembe AB, Campbell O, Hanson C, Tripathi V, Wong KL, et al. Caesarean section provision and readiness in Tanzania: analysis of cross-sectional surveys of women and health facilities over time. BMJ open. 2018;8:e024216.

9. Nahayo B, Olorunfemi G, Ndayishimye S, Nsanzabera C. Prevalence and factors associated with caesarean section among Tanzanian women of reproductive age: evidence from the 2022 Tanzania demographic and health survey data. BMC Public Health. 2025;25:794.

10. Nilsen C, Østbye T, Daltveit AK, Mmbaga BT, Sandøy IF. Trends in and socio-demographic factors associated with caesarean section at a Tanzanian referral hospital, 2000 to 2013. International journal for equity in health. 2014;13:1–11.

11. Sung S, Mikes BA, Martingano DJ, Mahdy H. Cesarean Delivery. StatPearls [Internet]. Treasure Island (FL): StatPearls Publishing; 2025 [cited 2025 Apr 17]. Available from: http://www.ncbi.nlm.nih.gov/books/NBK546707/

12. Bubpawong S, Nuampa S, Ratintorn A, Ruchob R. Multi-level Factors Influencing Caesarean Section Preferences among Women in Low- and Middle-Income Countries: A Systematic Review. Midwifery. 2025;104423.

13. Tarimo CS, Mahande MJ, Obure J. Prevalence and risk factors for caesarean delivery following labor induction at a tertiary hospital in North Tanzania: a retrospective cohort study (2000–2015). BMC Pregnancy and Childbirth. 2020;20:173.

14. Binyaruka P, Mori AT. Economic consequences of caesarean section delivery: evidence from a household survey in Tanzania. BMC Health Services Research. 2021;21:1367.

15. Idris IM, Menghisteab S. Cesarean section delivery rates, determinants, and indications: a retrospective study in Dekemhare Hospital. Global Reproductive Health. 2022;7:e56.

16. The United Republic of Tanzania (URT), Ministry of Finance and Planning, Tanzania, National Bureau of Statistics and President’s Office - Finance and Planning, Office of the, Chief Government Statistician, Zanzibar, Chief Government Statistician, Zanzibar. The 2022 Population and Housing Census: Administrative Units Population Distribution Report; Tanzania Zanzibar, [Internet]. The United Republic of Tanzania (URT); 2022. Available from: https://sensa.nbs.go.tz/publication/volume1c.pdf

17. Ministry of Health (MoH) [Tanzania Mainland], Ministry of Health (MoH) [Zanzibar], National Bureau of Statistics (NBS), Office of the Chief Government Statistician (OCGS), and ICF. Tanzania Demographicand Health Survey and Malaria Indicator Survey 2022 Key Indicators Report. Dodoma, Rockville: MoH, NBS, OCGS, and ICF; 2023.

18. Stata. Statistical software for data science [Internet]. [cited 2025 Apr 4]. Available from: https://www.stata.com/

19. WHO. WHO Statement on Caesarean Section Rates [Internet]. WHO; 2015. Available from: who.int/publicationsbitstream/handle/10665/161442/WHO_RHR_15.02_eng.pdf?sequence=1

20. Tull K. Maternal and sexual reproductive health situation in Tanzania. 2020;

21. Kibe PM, Mbuthia GW, Shikuku DN, Akoth C, Oguta JO, Ng’ang’a L, et al. Prevalence and factors associated with caesarean section in Rwanda: a trend analysis of Rwanda demographic and health survey 2000 to 2019–20. BMC pregnancy and childbirth. 2022;22:410.

22. Yaya S, Uthman OA, Amouzou A, Bishwajit G. Disparities in caesarean section prevalence and determinants across sub-Saharan Africa countries. Global Health Research and Policy. 2018;3:19.

23. Islam MdA, Sathi NJ, Hossain MdT, Jabbar A, Renzaho AMN, Islam SMS. Caesarean delivery and its association with educational attainment, wealth index, and place of residence in Sub-Saharan Africa: a meta-analysis. Scientific Reports. 2022;12:5554.

24. Souza J, Gülmezoglu A, Lumbiganon P, Laopaiboon M, Carroli G, Fawole B, et al. Caesarean section without medical indications is associated with an increased risk of adverse short-term maternal outcomes: the 2004-2008 WHO Global Survey on Maternal and Perinatal Health. BMC Medicine. 2010;8:71.

25. Boerma T, Ronsmans C, Melesse DY, Barros AJ, Barros FC, Juan L, et al. Global epidemiology of use of and disparities in caesarean sections. The Lancet. 2018;392:1341–8.

26. Rydahl E, Declercq E, Juhl M, Maimburg RD. Cesarean section on a rise—Does advanced maternal age explain the increase? A population register-based study. PloS one. 2019;14:e0210655.

27. Alipour A, Hantoushzadeh S, Hessami K, Saleh M, Shariat M, Yazdizadeh B, et al. A global study of the association of cesarean rate and the role of socioeconomic status in neonatal mortality rate in the current century. BMC Pregnancy and Childbirth. 2022;22:821.

28. Fotso JC, Speizer IS, Mukiira C, Kizito P, Lumumba V. Closing the poor-rich gap in contraceptive use in urban Kenya: are family planning programs increasingly reaching the urban poor? International Journal for Equity in Health. 2013;12:71.

29. Ana Pilar Betran, Jiangfeng Ye, Ann-Beth Moller, João Paulo Souza, Jun Zhang. Trends and projections of caesarean section rates: global and regional estimates. BMJ Global Health. 2021;6:e005671.

30. Shibre G, Zegeye B, Ahinkorah BO, Keetile M, Yaya S. Magnitude and trends in socio-economic and geographic inequality in access to birth by cesarean section in Tanzania: evidence from five rounds of Tanzania demographic and health surveys (1996–2015). Archives of Public Health. 2020;78:80.

